# Understanding COVID-19 vaccine hesitancy in Pakistan: The paradigm of Confidence, Convenience, and Complacency; A Cross-sectional study

**DOI:** 10.1101/2021.07.12.21260409

**Authors:** Naveen Siddique Sheikh, Mumtaz Touseef, Riddah Sultan, Kanwal Hassan Cheema, Sidra Shafiq Cheema, Afia Sarwar, Haniya Zainab Siddique

## Abstract

**Background and objectives:** Vaccine hesitancy is a big obstacle for vaccination programs, as is anticipated for the COVID-19 vaccination program, resulting in low uptake of vaccines thereby hindering the process of reaching herd immunity. Bearing this in mind the current study was aimed to explore the determinants of vaccine hesitancy amongst the Pakistani population.

**Methodology:** A cross-sectional study was carried out from November 2020 to March 2021. The conceptual framework of the study was based on the 3Cs (Confidence, Convenience, Complacency) model. The google-forms-based questionnaire was disseminated amongst the general population. Data collected were entered into SPSS version 26 and analyzed.

**Results:** Of the 421 participants, 68.4% were women. Non-healthcare workers were 55.8% of respondents. Of vaccine-hesitant individuals, 26.13% reported they were very unlikely to get vaccinated. The vaccine was not safe as it came out too fast was agreed upon by 12.6% of individuals, 50.6% were worried about experiencing side-effects, 18% believed the vaccine will not offer protection and 5.9% believed the vaccine would cause death. Low Practice of SOP in non-Healthcare workers was the strongest contributor to vaccine hesitancy (OR: 5.338, p=0.040, 95% CI: 1.082-26.330) followed by High complacency (p=0.026) and Moderate Complacency (OR: 0.212, p=0.007, 95% CI: 0.069-0.654) towards COVID-19 vaccination. In Healthcare workers the strongest contributor to vaccine hesitancy was having a Moderate Confidence (OR: 0.323, p=0.042, 95% CI: 0.109-0.958) in the vaccine followed by Moderate Convenience (OR: 0.304, p=0.049, 95% CI: 0.093-0.993) for vaccination

**Conclusion:** Campaigning and communication strategies to reaffirm confidence in the COVID-19 vaccine and educating the general population about the vaccine could lead to increased perception of vaccine safety and effectiveness thereby restoring confidence in vaccine and decreasing vaccine hesitancy. Likewise, working to increase vaccine convenience and decreasing complacency towards the COVID-19 vaccine would translate into high vaccine uptake.

## Introduction

‘Vaccine hesitancy refers to delayed acceptance or refusal of vaccination, despite resource availability(1)^]^. This well-known phenomenon endorsed into the present day is as old as the vaccine themselves(2), dating back to resistance programs against the mandated smallpox vaccination initiative in the mid-1800s(3). Consequently, over the years due to this phenomenon, vaccine-preventable diseases (VPD) the likes of measles, pneumococcal disease, pertussis, and poliomyelitis have resurfaced(4). The most serious instance of this was cited in the 2003-04 Northern Nigeria boycott of the polio vaccine, which led to the incidence of newer cases in the country(5).

A year has elapsed since the index case of the novel severe acute respiratory syndrome coronavirus 2 (SARS-CoV-2) was reported(6). On 31^st^ January 2020, World Health Organisation (WHO) declared a global health emergency and the coronavirus disease 2019 (Covid-19) was labeled a pandemic on 11^th^ March 2020(7). Since then, as of 26th January 2021, there have been 99 million cases of infection(8), 2.1 million deaths(9), and an economic loss of $3.7 trillion in earnings to workers around the world due to COVID-19(10).

The ongoing pandemic can be mitigated by an essential tool, an efficacious vaccine(s), which can reduce disease incidence, prevalence, new hospitalizations, and intensive care demand(11). The Pfizer-BioNTech’s (BNT162b2) and Moderna (mRNA-1273) mRNA vaccines were approved for emergency use by WHO in December 2020, giving hope for the resumption of normalcy(12). Experts estimated that herd immunity would be achieved if 70% of the population is immune to COVID-19(13). However, vaccine hesitancy amongst the general public can be a significant roadblock towards ensuring adequate vaccination uptake and achieving herd immunity. Vaccine hesitancy is becoming an impediment towards VPD prevention strategies, similar is anticipated for the forthcoming SARS-CoV-2 vaccine(14), consequently curbing the pandemic would become difficult with resistance to a prospective vaccine program.

Vaccine hesitancy is attributable to the ‘3Cs’ model which comprises confidence, complacency, and convenience(15). A lack of confidence in the vaccine safety, efficacy, or its delivery system; complacency due to a perceived low risk from VPD and vaccine inconvenience due to an inability to afford, hampers the success of vaccination campaigns(16). Further evaluation of the vaccine hesitancy reveals that public approval of vaccination is not motivated by empirical evidence-based medicine or economic data alone, but is rather driven by a combination of complex variables like political, psychological, technical, and sociocultural, all of which must be recognized and taken into consideration by policymakers and decision-makers(17). In addition, conspiracy beliefs result in vaccine hesitancy by undermining the trust in government bodies, healthcare workers, and pharmaceutical industries despite knowing their negative implications on human health behavior(18,19). Skepticism around the vaccines being adequately tested for safety, coming out too fast, and being registered in less than a year perpetuated the social media, thereby mediating low acceptance of the potential vaccine(20). Beliefs like vaccine causes infertility and is a means to stop population growth, is designed for electronic tattooing or microchipping individuals to achieve global surveillance and falsely asserting that vaccine causes autism, eventually lead to low vaccine uptake by the general population by undermining public confidence in vaccines and negatively impacting their attitude towards vaccination(21).

Public confidence and trust in vaccinations are highly variable. Building a group’s trust in vaccines requires that one understands their perception of vaccine and vaccine-associated risks or side effects, their socioeconomic standing, political stance, and religious affiliation. Although providing factually precise, accurate, scientifically sound evidence on the risk-benefit ratios of vaccines is of paramount importance, it is not sufficient to bridge the gap between present levels of confidence afforded by the public to vaccines and levels of trust required to ensure sufficient and continued vaccine coverage(4). In light of all this, the present study was planned to evaluate the determinants of COVID-19 vaccine hesitancy amongst the general Pakistani population. To also gauge their Willingness-to-pay (WTP)(22), thereby, ascertaining the amount they are inclined to allocate to vaccine technology.

## Methodology

A cross-sectional study was conducted from November 2020 to March 2021. Participants for the study were recruited from 23^rd^ January to 31^st^ January 2021 through the convenience sampling technique. Research questionnaire **S1 Appendix** constructed on google-forms was disseminated via online social media platforms (Facebook, WhatsApp, and Gmail) amongst the general population. Inclusion criteria were individuals above the age of 18 and residents of Pakistan. Exclusion criteria were minors and individuals residing outside of Pakistan. Items in the questionnaire were based on previous literature(23); some were modified taking into account the general population of the country. Informed electronic consent was the first part of the questionnaire. Participants were explained the voluntary nature of their participation, thereafter their consent was sought prior to filling out the questionnaire and collecting data (https://doi.org/10.6084/m9.figshare.14814882). The research was approved by the Institutional Review Board of CMH Lahore Medical College and Institute of Dentistry (Case#539/ERC/CMH/LMC).

### Sample Size

Sample size was calculated to be 385 using the formula:

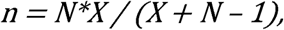

where,

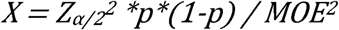

Zα/2 = critical value of the Normal distribution at α/2 (confidence level 95%, α= 0.05 and the critical value is 1.96)

MOE= margin of error= 5% p= sample proportion= 50%

N= population size= 220 million for Pakistan.

### Questionnaire instrument

The questionnaire comprised of four parts; demographics, a knowledge scale, and 2 sections exploring the beliefs, myths, and attitudes towards the COVID-19 vaccine and alternate preventive measures.

### Measures

Socio-demographic section recorded their gender, age group, education status, marital status, employment status, healthcare worker status, chronic disease status, and disease type. The Knowledge scale comprised of 10 questions. This tool had yes/no/I am not sure and multiple-choice questions. ‘Yes’ was scored as 1 point, ‘No’ and ‘I am not sure’ were scored 0 points. Those with multiple choice answers had a correct answer scored at 1 and incorrect answers scored at 0. Two Likert scale items (5 point Likert scale) were also employed. The tool included questions, about knowledge of vaccine existence, government’s initial plan of vaccination, re-infection, vaccination helping decrease spread of coronavirus infection, the demographic to which it could be given (children, pregnant and breastfeeding women), dose count, vaccine effectivity, and route of administration, framed in an approach similar to previous studies (23)(24)(25)(26). The score was measured by calculating the mean score of the 10 items. The lowest possible score was 2 and the highest possible score was 18.

A separate scale to explore the perceptions of respondents, on a series of items about COVID-19 infection (n = 4), a potential COVID-19 vaccination (n = 30), and COVID-19 vaccination cost (n=2) to establish the Willingness-To-pay, was used. Respondents rated the perception statements on a five-point Likert scale (1–5) from “strongly disagree” to “strongly agree”. Statements employed in the tool measured the theoretical constructs such as imagining themselves as being in a high-risk group, advantages of a potential COVID-19 vaccine, subjective norms, factors influencing their decision to vaccinate, behavioral control, myths, and beliefs about the vaccine, confidence in the Government and religious heads(23). These statements also probed respondents’ views on the vaccine enabling life to return to “normal,” and them being expected to adhere to the protocol for social distancing and other limitations for COVID-19 once vaccinated, along with items gauging their acknowledgment and practice of other preventive measures. Respondents were also inquired if they would vaccinate if their employer seeks proof of vaccination or they needed the proof for travel(23). These questionnaire items were computed to form 4 scales of Confidence, Convenience, Complacency (i.e 3C indicator), and SOP Practice as shown in **Table 2**. Mean scores were calculated and categorized on basis of the three percentiles i.e 33^rd^, 66^th^, 100^th^ percentile. The three groups were Low (below the 33^rd^ percentile), Moderate (between the 34^th^ and 66^th^ percentile), and High (above the 67^th^ percentile). A few items were calculated in the inverted sense to remain consistent with the direction of the indicator.

Finally, vaccination intention was also inquired (‘Yes’-intends to get vaccinated and ‘No’-does not intend to get vaccinated). The dichotomized response was used in the binary logistic regression model as the dependent variable with the demographics and the scales to find predictors of vaccine hesitancy. The data collected was analyzed using Statistical Package for the Social Sciences (SPSS) version 26. Data were analyzed for descriptive statistic analysis (means, standard deviations, frequency, and percentages) and inferential analysis (Independent sample *t*-test & Binary logistic regression). p-value ≤ 0.05 was considered statistically significant

## Results

The questionnaire was disseminated to 427 participants, out of which 421 completed it (98.5% response rate) by agreeing to the informed consent at the start of the questionnaire. Form for six participants was closed and submitted without being filled as they clicked disagree to the consent to fill. Women were 68.4% and men were 31.6%. The age group 20-30 years had the highest amount of respondents 70.3%. Of the 421 participants, 55.8% were not health care workers (HCW; doctors, nurses, technicians, paramedics, etc) while 44.2% were HCW. **Table 1** reflects the socio-demographic features of the respondents.

**Table 1:**
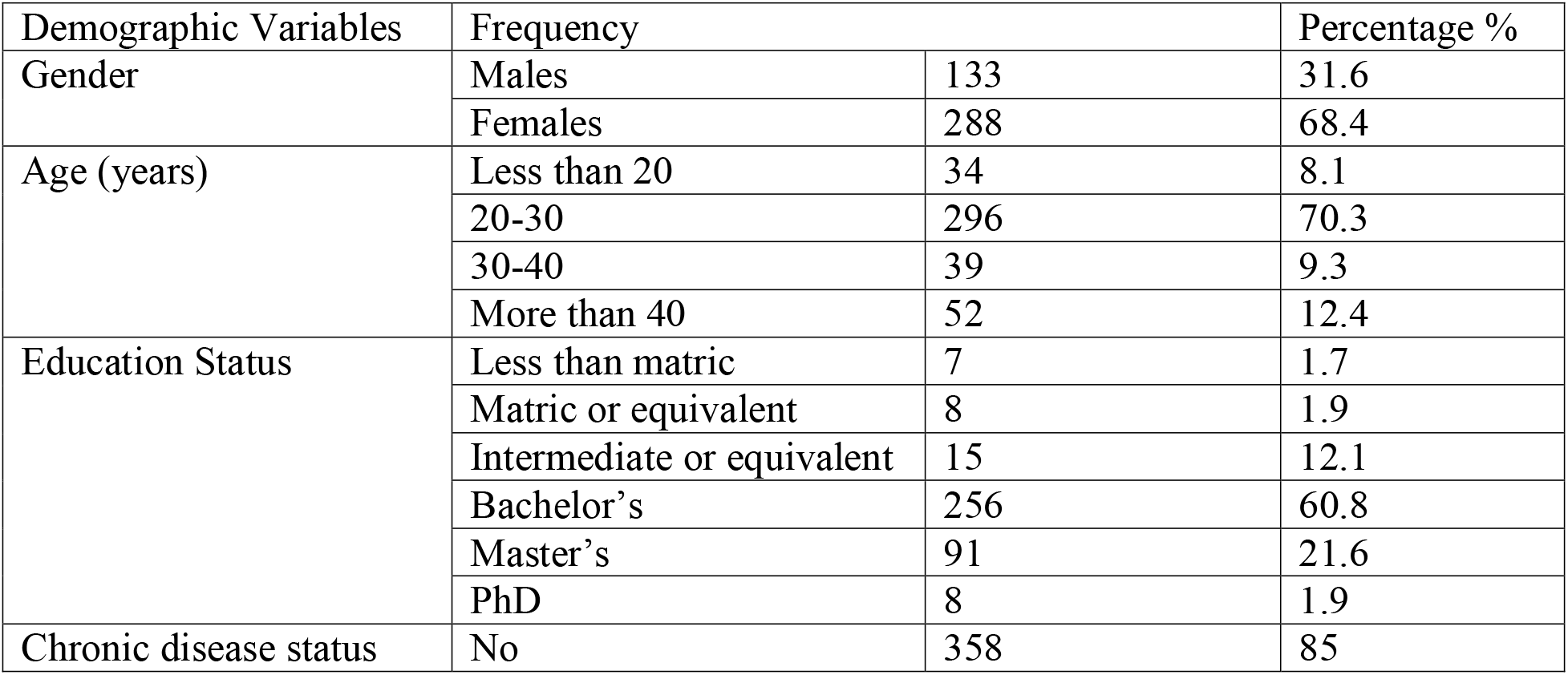
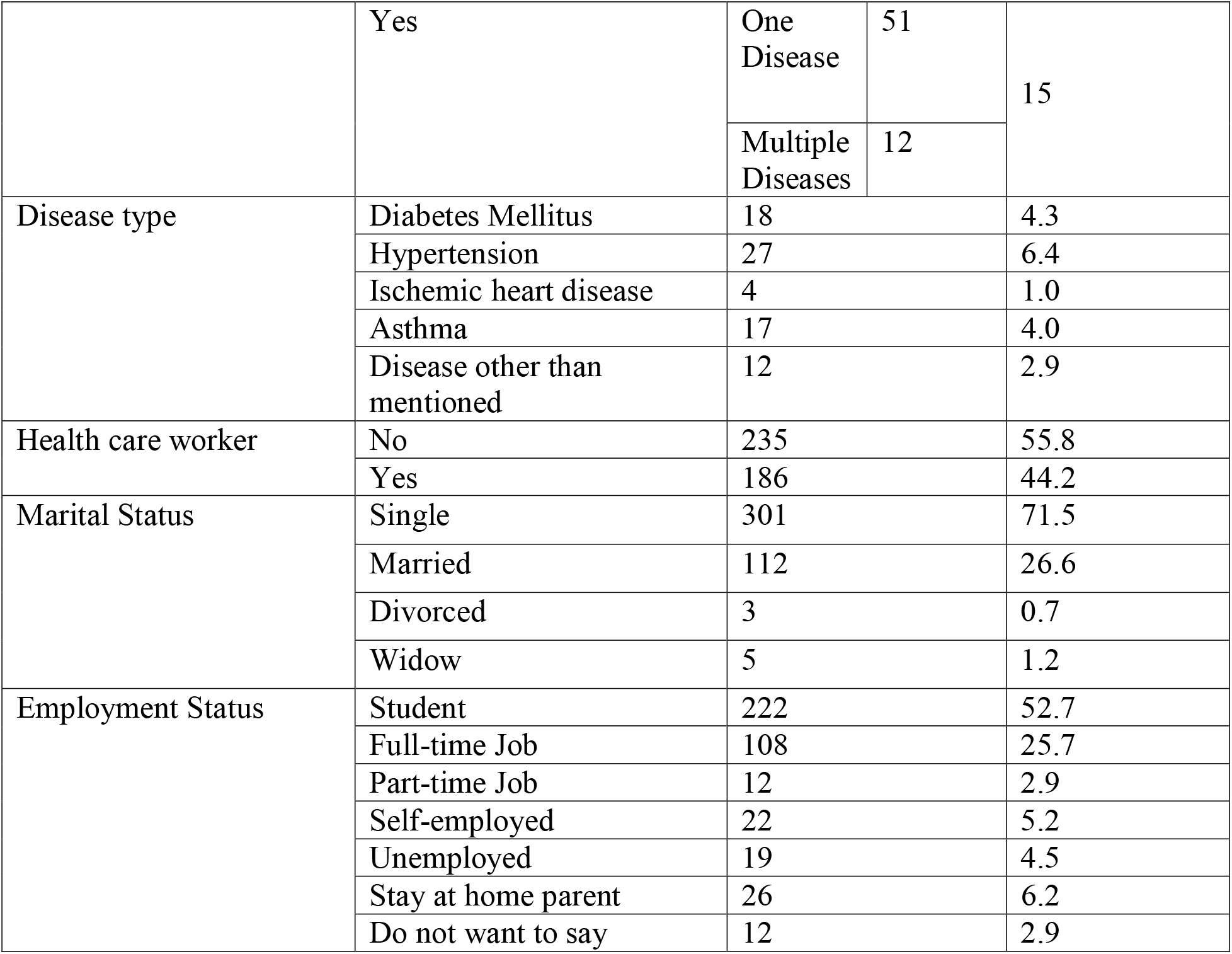
Socio-Demographic Details of Participants (N=421)

| Demographic Variables | Frequency |  | Percentage % |
| --- | --- | --- | --- |
| Gender | Males | 133 | 31.6 |
|  | Females | 288 | 68.4 |
| Age (years) | Less than 20 | 34 | 8.1 |
|  | 20-30 | 296 | 70.3 |
|  | 30-40 | 39 | 9.3 |
|  | More than 40 | 52 | 12.4 |
| Education Status | Less than matric | 7 | 1.7 |
|  | Matric or equivalent | 8 | 1.9 |
|  | Intermediate or equivalent | 15 | 12.1 |
|  | Bachelor's | 256 | 60.8 |
|  | Master's | 91 | 21.6 |
|  | PhD | 8 | 1.9 |
| Chronic disease status | No | 358 | 85 |

|  |  |  |  |  |
| --- | --- | --- | --- | --- |
|  | Yes | One Disease | 51 | 15 |
|  |  | Multiple Diseases | 12 |  |
| Disease type | Diabetes Mellitus | 18 |  | 4.3 |
|  | Hypertension | 27 |  | 6.4 |
|  | Ischemic heart disease | 4 |  | 1.0 |
|  | Asthma | 17 |  | 4.0 |
|  | Disease other than mentioned | 12 |  | 2.9 |
| Health care worker | No | 235 |  | 55.8 |
|  | Yes | 186 |  | 44.2 |
| Marital Status | Single | 301 |  | 71.5 |
|  | Married | 112 |  | 26.6 |
|  | Divorced | 3 |  | 0.7 |
|  | Widow | 5 |  | 1.2 |
| Employment Status | Student | 222 |  | 52.7 |
|  | Full-time Job | 108 |  | 25.7 |
|  | Part-time Job | 12 |  | 2.9 |
|  | Self-employed | 22 |  | 5.2 |
|  | Unemployed | 19 |  | 4.5 |
|  | Stay at home parent | 26 |  | 6.2 |
|  | Do not want to say | 12 |  | 2.9 |

Descriptive statistics for the items on the questionnaire are mentioned in **Table 2**. It also highlights how the 3C indicator was constructed. Statements other than the ones mentioned in the table used in the construction of the 3C indicator were the government’s initial plan for vaccination and vaccine effectivity in Confidence indicator; the existence of a vaccine, the vaccination being an almost pain-free procedure, the vaccine being able to decrease the spread of infection and whether a person could be re-infected with COVID-19 in Complacency indicator; and route of administration along with vaccine being dose-based in the Convenience indicator.

**Table 2:**
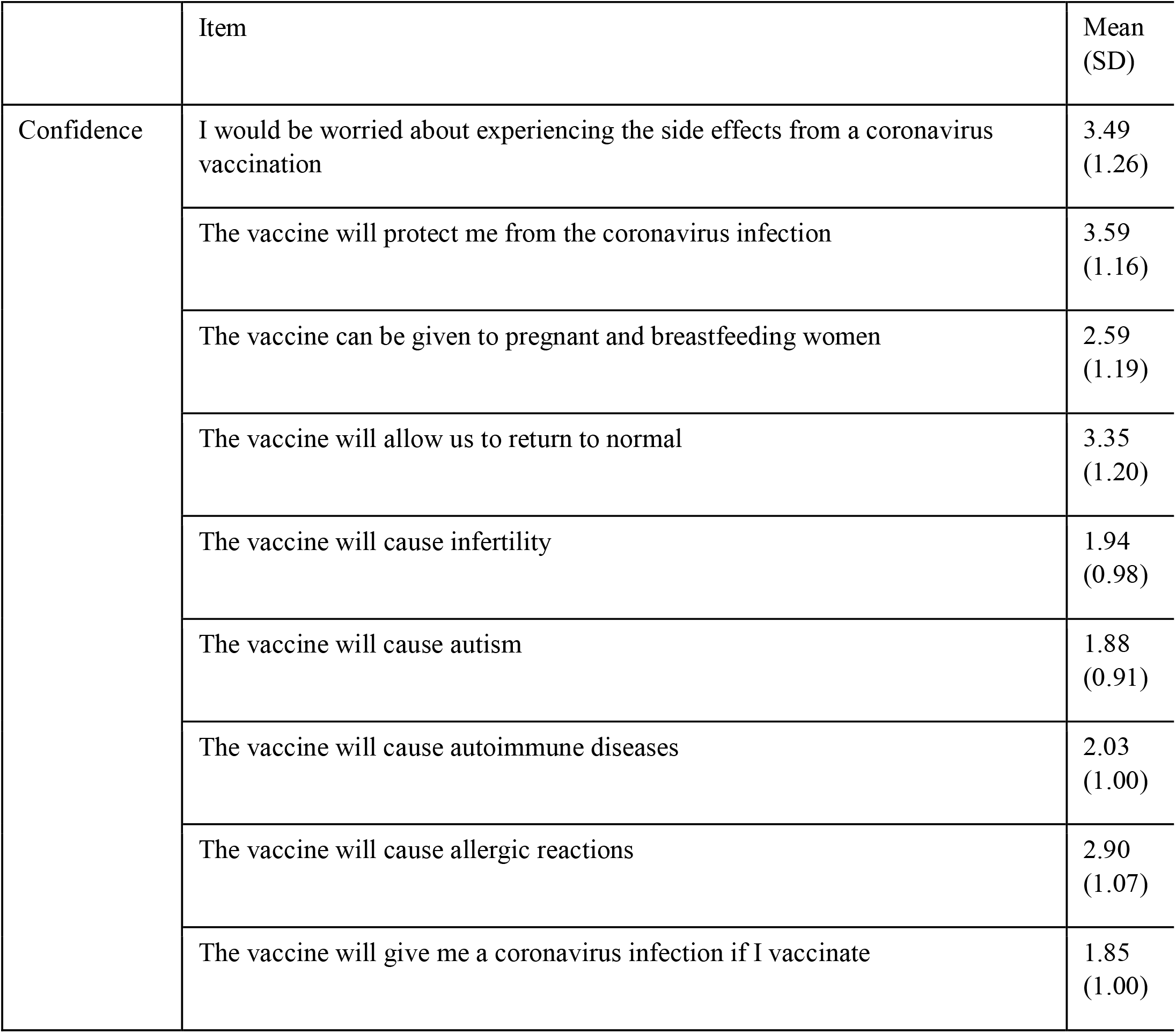
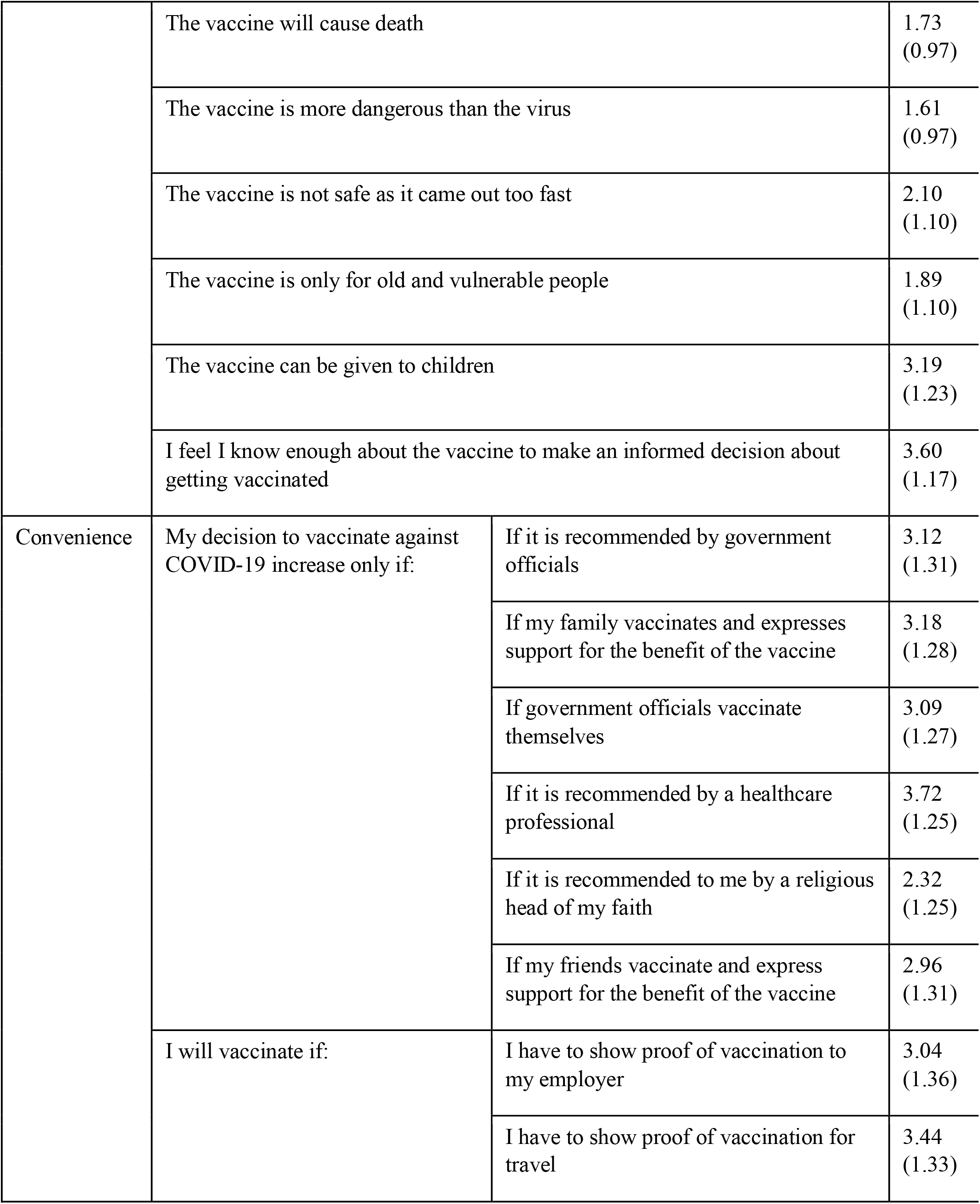
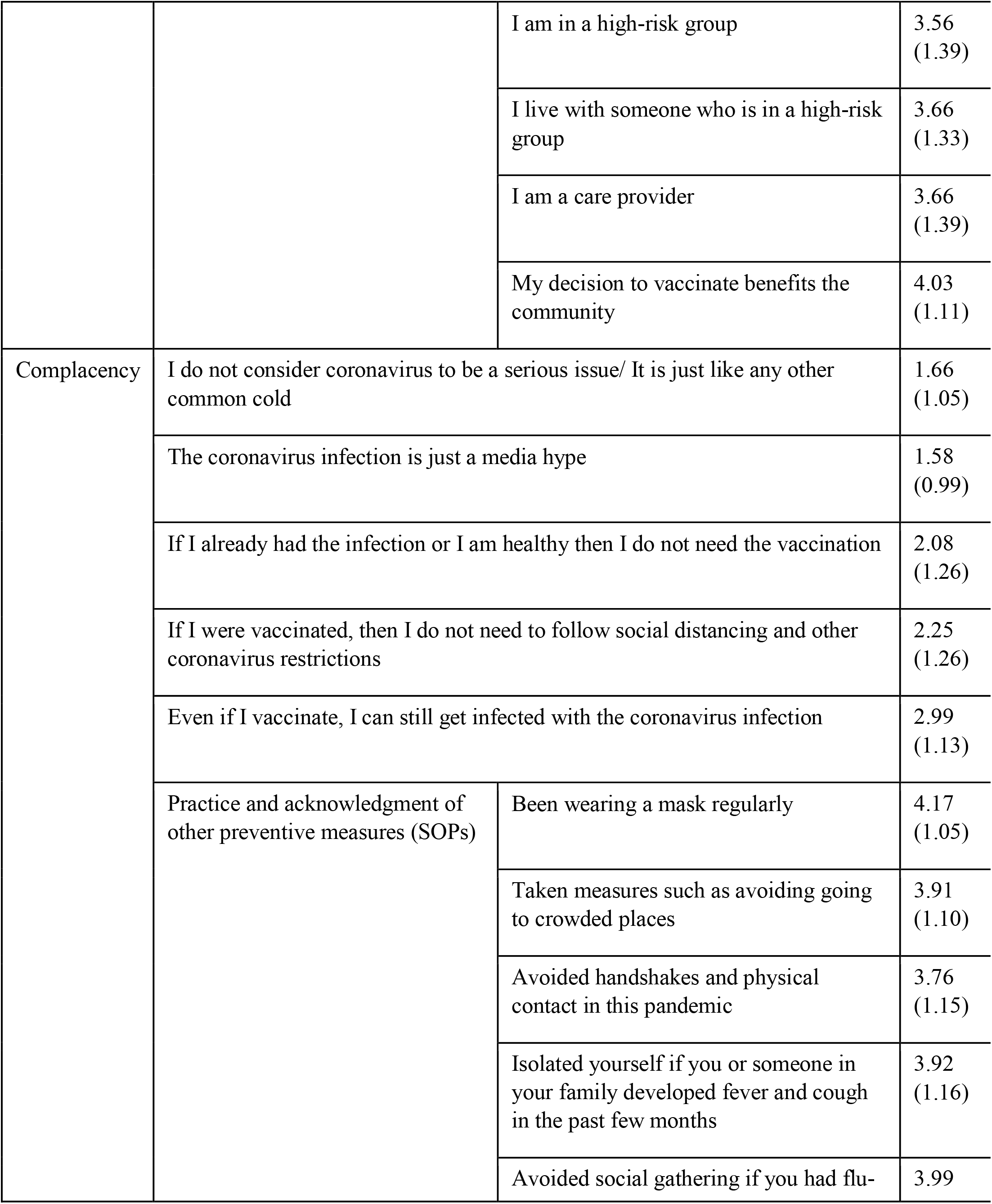
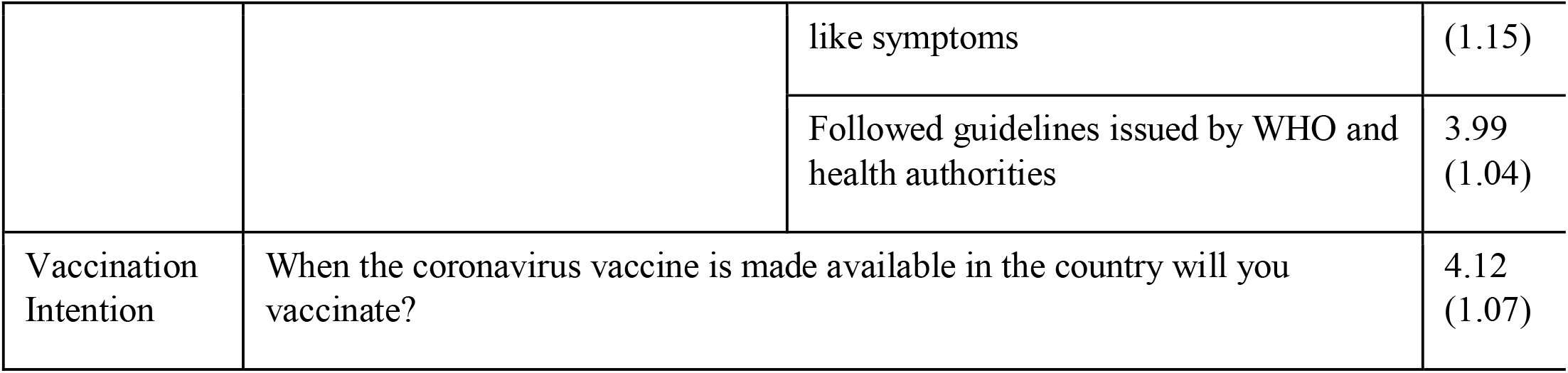
Descriptive statistics of continuous items evaluating Confidence, Complacency, Convenience, Practice of SOPS, and vaccination intention for COVID-19. Data are mean (standard deviation) on a 1–5 numerical rating scale (1 = strongly disagree, 5 = strongly agree).

Statements gauging the beliefs of people regarding the vaccine revealed that 14% agreed that if they were healthy or previously infected then they do not need the vaccination, 20% agreed that there is no need for social distancing once vaccinated, 18% agreed that the vaccine will not offer them protection, 28.8% agreed that vaccination will cause allergic reactions, 12.6% believed that the vaccine is not safe as it came out too fast and 45% disagreed that the vaccine could be given to pregnant/breastfeeding women. Fear of experiencing side-effects had 50.6% agree to the statement. What was worrisome was that a staggering 24% disagreed that they would not vaccinate if they were in a high-risk group and 20% disagreed to vaccinate even if they lived with someone in a high-risk group. Even if they were a care provider, 21.7% chose to disagree with vaccinating. Despite all this, 73% did however agree that the decision to vaccinate will benefit the community. When asked if they were aware that the state had an initial vaccination strategy for the population at high risk, healthcare workers, public health workers, and people with chronic diseases, only 58.7% of the non-healthcare workers (non-HCW) responded with a ‘Yes’ while 81.7% of the healthcare workers (HCW) responded with a ‘Yes’ (p-value 0.00). When asked about the dose count of the vaccine only 49.5% of the HCW responded correctly with 2 doses while only 34.0% of the non-HCW were aware of the correct response (p-value 0.001).

Two items in the questionnaire evaluated the Willingness-to-pay of the respondents. The first was who should cover the cost of the vaccine, and the second being that if you were to pay from pocket then how much are you willing to pay. The vaccine should be provided free of cost had 61.3% of the respondents choosing it, 28.7% chose that the vaccine be provided at a subsidized rate by the government while only 10% were willing to pay from pocket for the vaccine. On the amount of money that they were willing to pay, 27.3% chose less than 500 Pak rupees (PKR)($3.16), 31.8% chose 500-1000 PKR($3.16-6.32), 23% chose 1000-2000 PKR($6.32-12.64) while 17.8% were willing to pay more than 2000 PKR(>$12.64) for the COVID-19 vaccine. Inquiry about vaccination intention revealed, 73.87% reported they were likely to get vaccinated, 26.13% were unlikely to get vaccinated.

Scores were calculated for each scale and were then categorized into Low, Moderate, and High on a percentile basis. High scores on the Confidence and Convenience indicator translate into low vaccine hesitancy while high scores on the Complacency indicator translate into a high vaccine hesitancy. Mean scores of the respondents for each scale were calculated along with standard deviations highlighted in the **S1 Table**. An independent sample t-test was performed to see if the difference in means between healthcare workers and non-healthcare workers was true or due to chance. Results are tabulated in **Table 3** (also highlighting the percentage of respondents who scored in each category). The difference was significant in Knowledge and Confidence while non-significant for the remainder indicators.

**Table 3:**
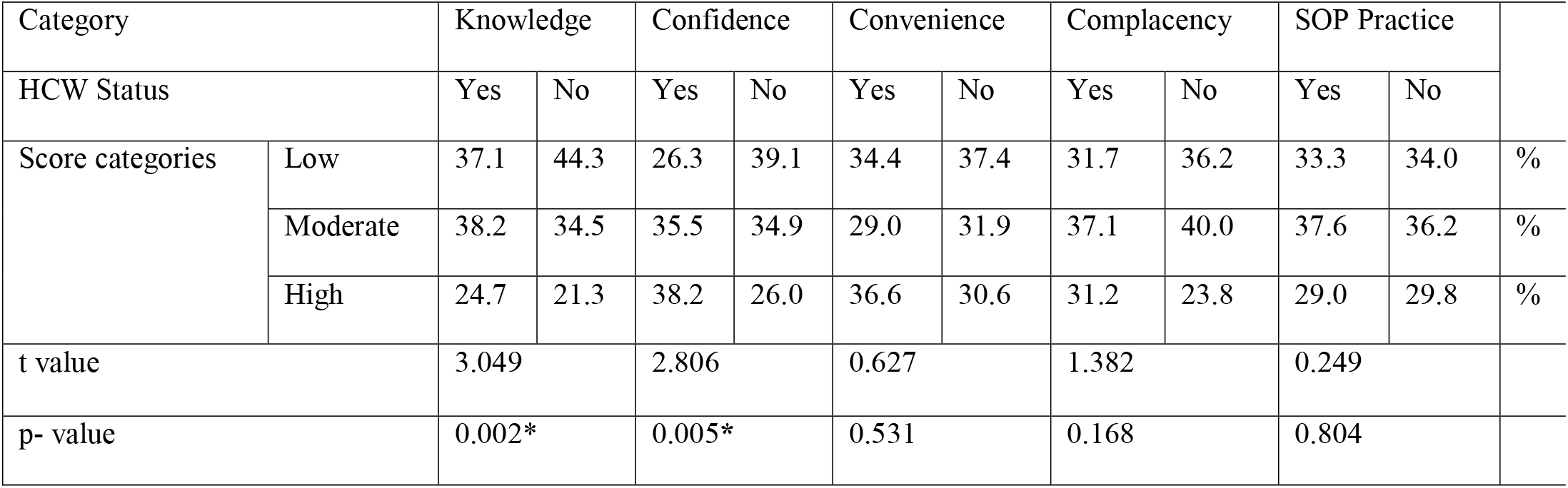
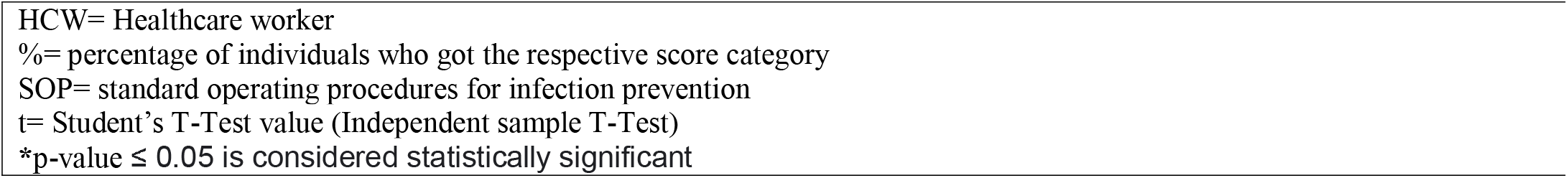
Independent Sample t-test for Knowledge, Confidence, Convenience, Complacency, and SOP Practice between Healthcare Workers (HCW) and Non-Healthcare Workers along with Score categorization for respective scales.

| Category |  | Knowledge |  | Confidence |  | Convenience |  | Complacency |  | SOP Practice |  |  |
| --- | --- | --- | --- | --- | --- | --- | --- | --- | --- | --- | --- | --- |
| HCW Status |  | Yes | No | Yes | No | Yes | No | Yes | No | Yes | No |  |
| Score categories | Low | 37.1 | 44.3 | 26.3 | 39.1 | 34.4 | 37.4 | 31.7 | 36.2 | 33.3 | 34.0 | % |
|  | Moderate | 38.2 | 34.5 | 35.5 | 34.9 | 29.0 | 31.9 | 37.1 | 40.0 | 37.6 | 36.2 | % |
|  | High | 24.7 | 21.3 | 38.2 | 26.0 | 36.6 | 30.6 | 31.2 | 23.8 | 29.0 | 29.8 | % |
| t value |  | 3.049 |  | 2.806 |  | 0.627 |  | 1.382 |  | 0.249 |  |  |
| p- value |  | 0.002* |  | 0.005* |  | 0.531 |  | 0.168 |  | 0.804 |  |  |

HCW= Healthcare worker %= percentage of individuals who got the respective score category SOP= standard operating procedures for infection prevention t= Student's T-Test value (Independent sample T-Test) \*p-value $\leq 0.05$ is considered statistically significant

A binary logistic regression analysis was performed to assess the relationship between Demographics, Knowledge, 3Cs indicator, SOP practice, and HCW status as predictors for vaccine hesitancy among the two groups. The model explained between 19.0% (Cox & Snell R square) and 28.2% (Nagelkerke R square) of the variance in Vaccine Hesitancy among HCW and between 16.3% (Cox & Snell R square) and 23.7% (Nagelkerke R square) for non-HCW. It correctly classified 76.3% of cases for HCW and 75.3% of cases for non-HCW. **Table 4** shows how each item made a statistically significant contribution to the model. Predicted probabilities were for membership of ‘not vaccinating’. Low Practice of SOP in non-HCW was the strongest contributor to vaccine hesitancy (OR: 5.338, p=0.040, 95% CI: 1.082-26.330) followed by High complacency (p=0.026) and Moderate Complacency (OR: 0.212, p=0.007, 95% CI: 0.069-0.654) towards COVID-19 vaccination. In HCW the strongest contributor to vaccine hesitancy was having a Moderate Confidence (OR: 0.323, p=0.042, 95% CI: 0.109-0.958) in the vaccine followed by Moderate Convenience (OR: 0.304, p=0.049, 95% CI: 0.093-0.993) for vaccination.

**Table 4:**
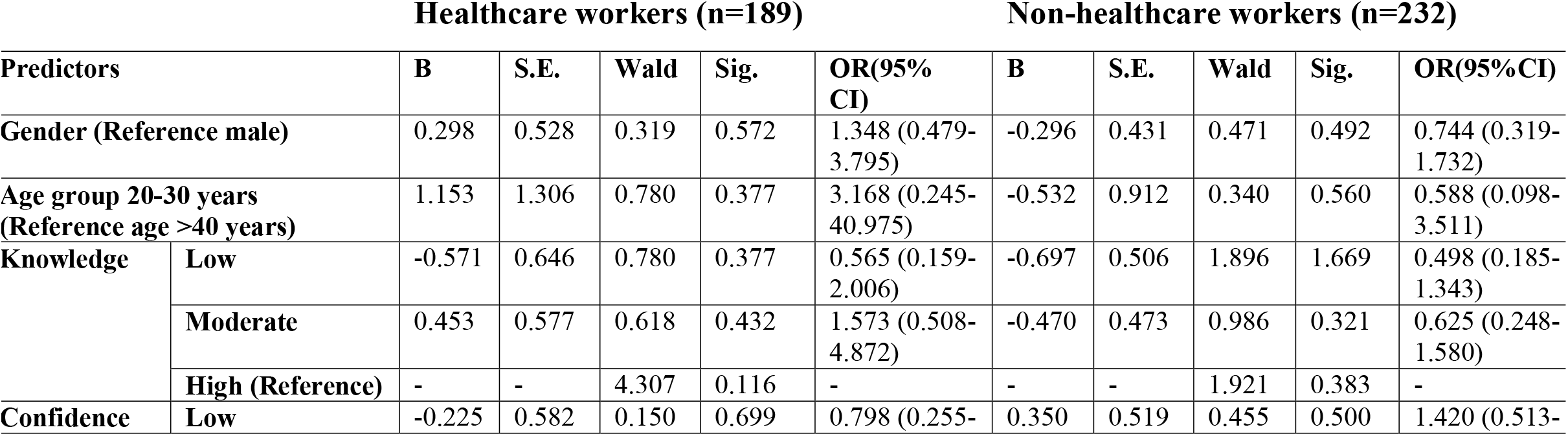
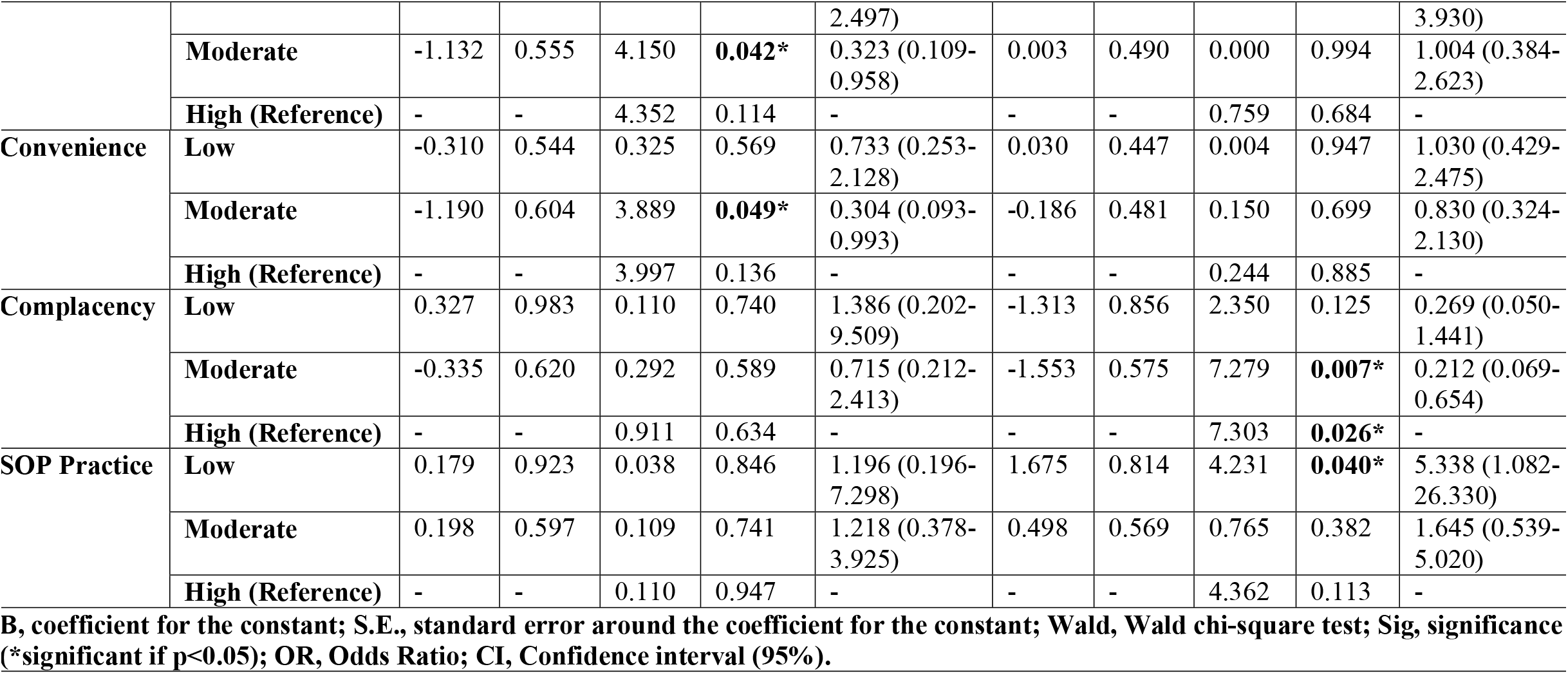
Binary logistic regression analysis predicting the likelihood of vaccine hesitancy amongst the respondents.

## Discussion

Vaccine hesitancy has a great role to play in the success of a vaccination program(27). Identifying the reasons for hesitancy and addressing them properly can help curb vaccine hesitancy thereby leading to the high uptake of vaccines and consequently reaching herd immunity faster. Our study resolved to find the determinants of vaccine hesitancy amongst the Pakistani population. The study revealed that 50.6% of respondents were worried about experiencing side-effects from the vaccine, a contributor to vaccine hesitancy, these findings are in resonance with a study published by the Centre for Economic Research in Pakistan (CERP) which reported that 54% of respondents to their study from Pakistan were worried about the safety of the vaccine(28). When compared to the study conducted in the United States of America which reported the count of individuals worried about the safety of the vaccine to be at 63.47%, our figures of 50.6% from Pakistan are significantly lower than theirs(29). Beliefs like vaccine will cause infertility (5.8% agreed), autism (3.1% agreed), autoimmune diseases (6.9% agreed), allergic reactions (28.8% agreed), death (5.9% agreed), is not safe as it came out too fast (12.6% agreed) and the vaccine is more dangerous than the virus (5% agreed) are unsurprising, they contributed to vaccine hesitancy and have been documented in previous literature(30). Such concerns need to be addressed by the health officials (physicians, public health workers) by having a healthy dialogue with the vaccine-hesitant people.

Of the respondents, 45% believed that the vaccine can not be given to pregnant/breastfeeding women, highlighting a lack of knowledge about the vaccine as The American College of Obstetricians and Gynaecologists(ACOG) recommends the vaccine for this group in guidelines with the Advisory Committee on Immunization Practices (ACIP)(31). Similarly, the finding that 43% agreed with administering the vaccine to children reflects a knowledge gap too, as CDC strictly prohibits the administration of the vaccine to individuals under the age of 16 years in the case of Pfizer and, 18 years in the case of the Moderna vaccine(32). Our research found that 61.7% of respondents agreed that they would vaccinate on the recommendation of a healthcare worker so having better physician recommendations is a good intervention to accentuate the success of an immunization program as also cited in previous literature(33).

Our findings revealed that Confidence, Convenience, and Complacency were statistically significant contributors to vaccine hesitancy, a finding documented in previous literature(34). Bearing this in mind, if policymakers work towards increasing confidence and convenience for the vaccine while decreasing complacency towards vaccine amongst the general population, they can subsequently variate the community’s intention to vaccinate significantly, this can be done by adopting tailored interventions for the context at hand concerning the different groups in the country, an approach supported in previous literature(35). Less adherence to COVID-19 health behaviors i.e low practice of SOPs was established as a contributor to vaccine hesitancy in our study, this finding is consistent with a previous study done in Australia which determined the same for COVID-19 vaccine hesitancy amongst the Australian population(36), similar was also reported in a study done in the US which cited that those who reported negative COVID-19 vaccination intentions had reduced odds of more frequent adherence to social distancing and wearing masks(37).

This research is not without its limitations and warrants enhancements. The data is cross-sectional making it difficult to disentangle causality of whether vaccine hesitancy is due to a lack of knowledge of vaccine and health information or due to the propensity to believe in conspiracy theories. The sample size is adequate, but it may not be truly representative of the entire Pakistani population, respondents were approached via convenience sampling through social networks of the data collectors, and considering that the authors belonged to only two provinces of the country, they were unable to collect equal responses from all the provinces. Furthermore, only 35% of the 224 million population of the country is urban and the country’s literacy rate is 60% with a mean of 5.2 years of schooling. While cellphone access is more than 50%, among youth, access to the internet is still however low (15%)(38). So the generalization of the results for the entire country should be done with caution. Vaccine hesitancy is a multifaceted problem, for which a better evaluation could be done via having a larger sample size with a longitudinal sampling approach and open-ended questions.

## Conclusion

Our study revealed that there was a significant knowledge gap regarding the vaccine. The belief in myths, like vaccine causes death, allergic reactions, and is more dangerous than the virus itself, was rampant. A lack of confidence in the vaccine, lack of convenience for the vaccine, and increased complacency were significant contributors to vaccine hesitancy. In light of the scale and scope of this major issue, all government and non-governmental health care departments must work together to restore trust in vaccines. The risk of vaccine-preventable disease(VPD), benefits of a vaccine for that VPD, and the risk-benefit ratio of the vaccine need to be discussed, supported by evidence-based medicine, with the hesitant people in a longitudinal, comprehensive, coherent, and in an unbiased transparent fashion to increase vaccination uptake(33). It is important to educate the public through mass outreach initiatives, awareness campaigns, and conferences to alleviate fear and uncertainty about the safety and efficacy of all vaccines and bridge the knowledge gap(39). Efforts to increase vaccine convenience and decrease complacency towards the COVID-19 vaccine would result in high vaccine uptake and a consequential faster herd immunity thereby decreasing the spread of infection and help in curbing the pandemic.

## Data Availability

Underlying data of the research is cited in the manuscript body.

https://doi.org/10.6084/m9.figshare.14814882

## Acknowledgments

We would like to thank Dr. Ahmed Waqas, Ph.D. fellow, Institute of Population Health, University of Liverpool, for his critical evaluation of research methodology, results, and manuscript revision. Our gratitude also goes to the study participants.

## Supplemental Information

**Table S1:**
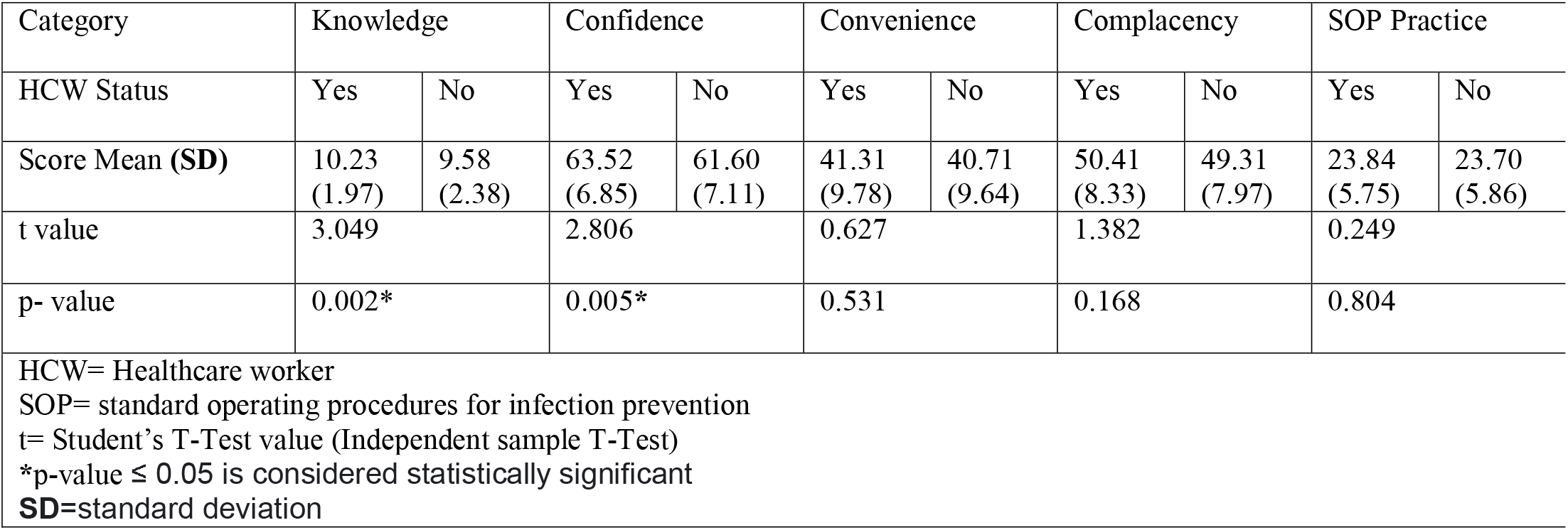
Independent Sample t-test for Knowledge, Confidence, Convenience, Complacency and SOP Practice between Healthcare Workers (HCW) and Non-Healthcare Workers.

## Notes

### Competing Interest Statement

The authors have declared no competing interest.

### Funding Statement

No external funding.

### Author Declarations

The research was approved by the Institutional Review Board of CMH Lahore Medical College and Institute of Dentistry, Lahore. (Case#539/ERC/CMH/LMC)

