## Supplementary material for "Understanding COVID-19 vaccine hesitancy in Pakistan: The paradigm of Confidence, Convenience, and Complacency; A Cross-sectional study": S1 Appendix

### COVID-19 Vaccine Questionnaire

Coronavirus is a rapidly spreading infection affecting millions worldwide. In an attempt to prevent further cases of infection and slow down the spread, scientists from all over the world are working towards developing a vaccine for the disease to prevent it. Through this study, the researchers intend to explore the knowledge, attitude, perception and willingness to vaccinate for the disease once the vaccine (questions pertain to Moderna/Pfizer/SinoPharm vaccine) is made available to the general public.

CONSENT FORM:
You are invited to participate in a web-based online survey on 'COVID-19 Vaccine'.

PARTICIPATION
Your participation in this survey is voluntary. You may refuse to take part in the research or exit the survey at any time without penalty. You are free to decline to answer any particular question you do not wish to answer for any reason.

BENEFITS
You will receive no direct benefits from participating in this research study. However, your responses may help us learn more about the improvements required.

RISKS
There are no foreseeable risks involved in participating in this study.

CONFIDENTIALITY
Your survey answers will be kept confidential and data will be stored in a password protected electronic format. Survey does not collect identifying information such as your name, email address, or IP address. Therefore, your responses will remain anonymous. No one will be able to identify you or your answers, and no one will know whether or not you participated in the study.

* Required

1. ELECTRONIC CONSENT: Please select your choice below. You may print a copy of this consent form for your records. Clicking on the “Agree” button indicates that: You have read the above information; You voluntarily agree to participate. *

*Mark only one oval.*

- ◯ Agree
- ◯ Disagree

#### Demographics

2. Gender?

*Mark only one oval.*

- ⬭ Male
- ⬭ Female
- ⬭ Other

3. Age (years)?

*Mark only one oval.*

- ⬭ Less than 20
- ⬭ 20-30
- ⬭ 31-40
- ⬭ More than 40

4. Education Level?

*Mark only one oval.*

- ⬭ Less than matric
- ⬭ Matric or equivalent
- ⬭ Intermediate or equivalent
- ⬭ Bachelor's
- ⬭ Master's
- ⬭ PhD

5. Are you a healthcare provider? (Doctor, nurse, technician, paramedic etc)

*Mark only one oval.*

- Yes
- No

6. Marital Status?

*Mark only one oval.*

- Single
- Married
- Divorced
- Widow

7. Employment?

*Mark only one oval.*

- Full-time job
- Part-time job
- Self-employed
- Unemployed but looking for a job
- Stay at home parent
- Student
- Do not want to disclose

8. Do you have a chronic disease?

*Check all that apply.*

- [ ] Diabetes (sugar)
- [ ] Hypertension (blood pressure)
- [ ] Heart disease
- [ ] Tuberculosis (TB)
- [ ] Asthma
- [ ] Disease other than listed above
- [ ] I do not have a chronic disease

#### Section 1

This section explores your knowledge of the disease and its vaccine

9. Do you know there is a vaccine for the coronavirus infection?

*Mark only one oval.*

- ( ) Yes
- ( ) No
- ( ) I am not sure

10. Do you know the route of administration of the vaccine?

*Mark only one oval.*

- ( ) Oral drops
- ( ) Tablet
- ( ) Injection in muscle
- ( ) IV infusion
- ( ) Syrup
- ( ) I am not sure

11. Do you know that the state has an initial vaccination strategy for population at high risk, healthcare workers, public health workers and people with chronic disease?

*Mark only one oval.*

- Yes
- No
- I am not sure

12. Vaccination is an almost pain-free procedure?

*Mark only one oval.*

- Yes
- No
- I am not sure

13. The coronavirus vaccine can help in decreasing the spread of coronavirus infection?

*Mark only one oval.*

- Yes
- No
- I am not sure

14. How effective is the coronavirus vaccine?

*Mark only one oval.*

- Not that effective
- Effective
- I am not sure

15. A person can be re-infected with the coronavirus?

*Mark only one oval.*

- Yes
- No
- I am not sure

16. Coronavirus vaccine is given in doses:

*Mark only one oval.*

- 1 dose
- 2 doses
- ≥3 doses
- I am not sure

#### Section 2

This section explores the different myths, beliefs and perspectives about the disease and its vaccination.

17. I do not consider coronavirus to be a serious issue/it is just like any other common cold.

*Mark only one oval.*

| | 1 | 2 | 3 | 4 | 5 | |
|---|---|---|---|---|---|---|
| Strongly Disagree | | | | | | Strongly Agree |

18. The coronavirus infection is just a media hype

*Mark only one oval.*

| | 1 | 2 | 3 | 4 | 5 | |
|---|---|---|---|---|---|---|
| Strongly Disagree | ◯ | ◯ | ◯ | ◯ | ◯ | Strongly Agree |

19. If I already had the infection or I am healthy then I do not need the vaccination

*Mark only one oval.*

| | 1 | 2 | 3 | 4 | 5 | |
|---|---|---|---|---|---|---|
| Strongly Disagree | ◯ | ◯ | ◯ | ◯ | ◯ | Strongly Agree |

20. I would be worried about experiencing the side effects from a coronavirus vaccination

*Mark only one oval.*

| | 1 | 2 | 3 | 4 | 5 | |
|---|---|---|---|---|---|---|
| Strongly Disagree | ◯ | ◯ | ◯ | ◯ | ◯ | Strongly Agree |

21. If I were vaccinated, then I think I do not need to follow social distancing and other coronavirus restrictions

*Mark only one oval.*

| | 1 | 2 | 3 | 4 | 5 | |
|---|---|---|---|---|---|---|
| Strongly Disagree | ◯ | ◯ | ◯ | ◯ | ◯ | Strongly Agree |

22. I FEEL THAT THE VACCINE: (Please indicate the extent to which you agree or disagree with the following statements by selecting a number between 1 and 5, where: 1 means 'strongly disagree' and 5 means ' strongly agree').

*Mark only one oval per row.*

| | 1 | 2 | 3 | 4 | 5 |
|---|---|---|---|---|---|
| Will protect me from the coronavirus infection | ◯ | ◯ | ◯ | ◯ | ◯ |
| Vaccine can be given to pregnant and breastfeeding women | ◯ | ◯ | ◯ | ◯ | ◯ |
| Vaccine will allow us to return to normal | ◯ | ◯ | ◯ | ◯ | ◯ |
| Will cause infertility | ◯ | ◯ | ◯ | ◯ | ◯ |
| Will cause autism | ◯ | ◯ | ◯ | ◯ | ◯ |
| Will cause autoimmune diseases | ◯ | ◯ | ◯ | ◯ | ◯ |
| Will cause allergic reactions | ◯ | ◯ | ◯ | ◯ | ◯ |
| Will give me a coronavirus infection if I vaccinate | ◯ | ◯ | ◯ | ◯ | ◯ |
| Will cause death | ◯ | ◯ | ◯ | ◯ | ◯ |
| Vaccine is more dangerous than the virus | ◯ | ◯ | ◯ | ◯ | ◯ |
| Vaccine is not safe as it came out too fast | ◯ | ◯ | ◯ | ◯ | ◯ |
| Vaccine is only for old and vulnerable people | ◯ | ◯ | ◯ | ◯ | ◯ |
| Vaccine can be given to children | ◯ | ◯ | ◯ | ◯ | ◯ |
| Even if I vaccinate, I can still get infected with coronavirus infection | ◯ | ◯ | ◯ | ◯ | ◯ |

23. My decision to vaccinate against COVID-19 increases ONLY IF: (Please indicate the extent to which you agree or disagree with the following statements by selecting a number between 1 and 5, where: 1 means 'strongly disagree' and 5 means ' strongly agree')

*Mark only one oval per row.*

| | 1 | 2 | 3 | 4 | 5 |
|---|---|---|---|---|---|
| If it is recommended by government officials | | | | | |
| If my family vaccinates and expresses support for the benefit of vaccine | | | | | |
| If government officials vaccinate | | | | | |
| If it is recommended by a healthcare professional (GP, nurse family doctor) | | | | | |
| If it is recommended to me by a religious head of my faith | | | | | |
| If my friends vaccinate and express support for the benefit of the vaccine | | | | | |

24. I feel I know enough about the vaccine to make an informed decision about getting vaccinated

*Mark only one oval.*

| | 1 | 2 | 3 | 4 | 5 | |
|---|---|---|---|---|---|---|
| Strongly Disagree | | | | | | Strongly Agree |

#### Section 3

This sections explores your practice regarding the virus and its vaccination

25. When the coronavirus vaccine is made available in the country, will you vaccinate?

*Mark only one oval.*

- Yes
- No

26. I will vaccinate if: (This question concerns the cost of vaccination, More than 1 option can be selected)

*Mark only one oval.*

- Vaccine is provided free of cost by the government
- Vaccine is provided at a subsidized rate by the government
- I have to pay for the vaccine from my pocket

27. If I have to pay for the vaccine, then I am comfortable paying (In Pak Rupees PKR):

*Mark only one oval.*

- Less than 500 PKR
- 500-1000 PKR
- 1000-2000 PKR
- More than 2000 PKR

28. I WILL VACCINATE IF: (Please indicate the extent to which you agree or disagree with the following statements by selecting a number between 1 and 5, where: 1 means 'strongly disagree' and 5 means ' strongly agree')

*Mark only one oval per row.*

| | 1 | 2 | 3 | 4 | 5 |
|---|---|---|---|---|---|
| I have to show proof of vaccination to my employer | | | | | |
| I have to show proof of vaccination for travel | | | | | |
| I am in a high risk group | | | | | |
| I live with someone who is in a high risk group | | | | | |
| I am a care provider | | | | | |
| My decision to vaccinate benefits the community | | | | | |

29. HAVE YOU: (Please indicate the extent to which you agree or disagree with the following statements by selecting a number between 1 and 5, where: 1 means 'strongly disagree' and 5 means ' strongly agree')

*Mark only one oval per row.*

| | 1 | 2 | 3 | 4 | 5 |
|---|---|---|---|---|---|
| Been wearing a mask regularly | ◯ | ◯ | ◯ | ◯ | ◯ |
| Taken measures such as avoided going to crowded places | ◯ | ◯ | ◯ | ◯ | ◯ |
| Avoided hand shakes and physical contact in this pandemic | ◯ | ◯ | ◯ | ◯ | ◯ |
| Isolated yourself if you or someone in your family developed fever and cough in the past few months | ◯ | ◯ | ◯ | ◯ | ◯ |
| Avoided social gatherings if you had flu like symptoms | ◯ | ◯ | ◯ | ◯ | ◯ |
| Followed guidelines issued by WHO and health authorities | ◯ | ◯ | ◯ | ◯ | ◯ |

30. Thank you for filling out the form. If you have any feedback for us, please leave it in the space below.

This content is neither created nor endorsed by Google.

Google Forms
