## Supplementary material for "Understanding COVID-19 vaccine hesitancy in Pakistan: The paradigm of Confidence, Convenience, and Complacency; A Cross-sectional study": S1 Table

**Supplemental Information:**

**S1 Table: Independent Sample t-test for** **Knowledge, Confidence, Convenience, Complacency and SOP Practice between Healthcare Workers (HCW) and Non-Healthcare Workers**

| Category | Knowledge | | Confidence | | Convenience | | Complacency | | SOP Practice | |
| --- | --- | --- | --- | --- | --- | --- | --- | --- | --- | --- |
| HCW Status | Yes | No | Yes | No | Yes | No | Yes | No | Yes | No |
| Score Mean **(SD)** | 10.23  (1.97) | 9.58  (2.38) | 63.52  (6.85) | 61.60  (7.11) | 41.31  (9.78) | 40.71  (9.64) | 50.41  (8.33) | 49.31  (7.97) | 23.84  (5.75) | 23.70  (5.86) |
| t value | 3.049 | | 2.806 | | 0.627 | | 1.382 | | 0.249 | |
| p- value | 0.002* | | 0.005***** | | 0.531 | | 0.168 | | 0.804 | |
| HCW= Healthcare worker  SOP= standard operating procedures for infection prevention  t= Student’s T-Test value (Independent sample T-Test)  *****p-value ≤ 0.05 is considered statistically significant  **SD**=standard deviation | | | | | | | | | | |
